# Caregivers’ Qualitative Insights on Trust, Resilience and Vaccine Hesitancy Shaping Child Health in Conflict-affected Northeast Nigeria

**DOI:** 10.1101/2025.09.18.25335857

**Authors:** Liliana Abreu, Pia Schrage, Gbadebo Collins Adeyanju, Rabiu Ibrahim Jalo, Aisha Aliyu Abulfathi, Musa Muhammad Bello, Aminatu Ayaba Kwaku, Muhammad Ibrahim Jalo, Ahmad Mahmud, Max Schaub

## Abstract

**Background:** The northeast of Nigeria, particularly the conflict-affected states of Borno, Yobe and Adamawa, has some of the highest under-five mortality rates in the world. Armed conflict, damaged health infrastructure and systemic poverty have significantly reduced access to healthcare. This study explores the intersection of health-seeking behaviour among caregivers of children under five with trust in health systems, exposure to violence, and vaccine hesitancy.

**Methods:** A qualitative study was conducted using nine focus group discussions (FGDs) with 72 caregivers living in conflict-affected communities. Participants were purposively selected, and the discussions explored the following topics: barriers to accessing healthcare; trust in health systems; the impact of conflict on health-seeking behaviour; and perceptions of childhood vaccinations. Data were analysed thematically using content analysis.

**Results:** Health-seeking behaviour was found to be shaped by a complex interplay of poverty, dysfunctional health infrastructure and deep-seated mistrust of governmental institutions, all of which were exacerbated by prolonged exposure to violence. Patriarchal norms played a central role in decision-making processes, often limiting women’s autonomy when it came to accessing care. Vaccine hesitancy was influenced by misinformation, knowledge gaps and limited community engagement. However, caregivers with access to reliable information sources, such as community networks and local media, showed more positive attitudes towards immunisation.

**Conclusions:** Efforts to improve maternal and child health in conflict-affected regions must prioritise trust-building, strengthening the health system in a culturally sensitive way, and implementing targeted communication strategies. Community engagement and resilience must be leveraged to reduce barriers to care and address vaccine hesitancy in fragile contexts.

## Introduction

Child health is a central priority in global health. It is a critical indicator of overall health equity, social justice, and the effectiveness and responsiveness of health systems worldwide. Specifically, under-five mortality is a major indicator of population health, development progress and the effectiveness of health interventions, particularly in low- and middle-income countries (LMICs). Despite significant global progress in child health, disparities remain, with regions affected by poverty, socio-political instability and conflict, bearing a disproportionate burden (1). In 2022, approximately 4.9 million children under the age of five died globally, with nearly half of these deaths occurring in sub-Saharan Africa (SSA) (2). This region continues to have high under-five mortality rates, mainly from preventable or treatable causes (3). Addressing these disparities is a priority for global health initiatives, however, achieving this goal is complicated due to high exposure to armed conflict and fragile health systems, which aggravate vulnerability and limit access to essential services such as vaccination or maternal and child health care. Nigeria’s under-five mortality rate remains one of the highest globally, far exceeding the Sustainable Development Goal (SDG) target of 25 deaths per 1,000 by 2030, which would require a reduction of at least 50% (4,5). Furthermore, under-five mortality rates in Nigeria vary widely across regions. In 2021, subnational data show striking contrasts, with under-five mortality rates of 52 per 1,000 in the south-west of the country compared with 253 per 1,000 in the north-west (6) when national average was 111 deaths per 1,000 live births (7).

One key driver of high child mortality rates in northeast Nigeria is the limited access to healthcare services, particularly within communities affected by ongoing conflict. For caregivers living in these regions under the constant threat of armed conflict, the decision to seek healthcare is influenced not only by perceived health needs or service availability, but also by the profound instability caused by ongoing violence (8). In fact, any health-seeking behaviour, defined as the actions taken by individuals to maintain, restore or improve health, is not solely an individual choice but results from a combination of social, cultural, psychological and structural determinants (9). In the northeast Nigerian context, where socio-political instability and conflict are widespread, caregivers’ health-related decisions may depend on factors such as their experiences of violence, trust in institutions, distance to health facilities, costs of treatment, or cultural practices – all of which can affect attitudes towards preventive health measures such as vaccination (10–14). The past years of conflict had a severe impact on the region’s physical health infrastructure. The extended violence, largely driven by the Boko Haram insurgency, has led to the destruction or dysfunction of over 40% of health facilities (16), significantly reducing the probability of maternal healthcare utilization, such as, antenatal care (ANC) visits (17). In addition, the well-known excessive use of force against civilians by security forces intensifies an already pervasive climate of fear and insecurity, creating additional and significant barriers that further discourage families from accessing essential health services (8,15).

### Exposure to armed conflict and vaccine hesitancy

Two significant indirect effects of armed conflict are the inhibition of immunization completion and maternal health service uptake (17,18). Armed conflict disrupts immunization services, leading to the resurgence and spread of preventable and previously eradicated illnesses, and erodes trust, exacerbating vaccine hesitancy among populations exposed to violence (11,19). This mistrust is particularly vulnerable when it comes to preventive services such as immunisation, as caregiver concerns are heightened by fears about vaccine safety, perceived government neglect, and socio-political rumours that fuel hesitancy. Overall, vaccine hesitancy is a particularly prominent issue in Nigeria (1,14,20). The World Health Organization defined vaccine hesitancy as "delay in accepting or refusing vaccines despite the availability of immunisation services", vaccine hesitancy is a multidimensional challenge influenced by historical, cultural and political contexts (21). In northeast Nigeria, vaccine hesitancy is rooted in local resistance and mistrust of government-led health initiatives, exemplified by the 2003 polio vaccine boycott, which has had a long lasting impact on attitudes towards vaccination in the region (22–25). Studies on vaccine hesitancy and child healthcare-seeking behaviors suggests that vaccine hesitancy and/or incomplete vaccinations are driven by a number of interrelated societal factors. A systematic review on reasons for non-vaccination among children and adolescents in SSA pointed towards lack of knowledge on vaccinations, time constraints for caregivers, lack of vaccinations and healthcare personnel, as well as caregiver’s beliefs, as major factors that contribute to incomplete vaccinations (26). Additionally, another three studies conducted in Nigeria pointed to pressure against vaccination either from the husband or partner or even for cultural or religious reasons (27,27–29). Gender dynamics in health-seeking behaviour and decision-making plays an important role. In a predominantly patriarchal society as Nigeria, women often have limited autonomy to make decisions about their own health and that of their children (22). Healthcare choices need to be approved by the male head of household despite known poor knowledge on maternal health (30). While mothers are the primary caretakers of children in Nigeria, their involvement in maternal and child care is usually sub-optimal (31) - the male head of household is the primary healthcare decision maker (22,32). Consequently, the patriarch’s level of education, employment status, household wealth and awareness of the importance of vaccinations directly informs the decision making process (33–37). In contrast, women with high decision making autonomy have significantly higher odds of ANC and childhood immunisation service uptake compared to those with low decision making autonomy (38–40). By exploring caregivers’ narratives, our study aims to contribute to the growing body of literature on health-seeking behaviour in conflict-affected settings and to inform culturally sensitive health interventions that address the specific needs and constraints faced by caregivers in such contexts.

## Objectives

This study aims to explore how health-seeking behaviour, exposure to conflict, trust and vaccine hesitancy intersect to shape caregivers’ decisions about accessing healthcare in northeast Nigeria. Through a qualitative approach, namely focus group discussions with caregivers, we sought to gain a deeper understanding of caregivers’ barriers and enablers to healthcare access in this region.

## Methods

### Setting and participants

This study used a qualitative research design to explore the relationship between health-seeking behaviour, trust in health systems and exposure to violence among caregivers of children under five in northeast Nigeria. The study was conducted in three conflict-affected states of northeast Nigeria (Borno, Yobe and Adamawa) in communities that have been significantly affected by ongoing armed violence. Participants included caregivers of children under five years of age, who were purposely selected based on their caregiving role and exposure to armed conflict (Table 1). Inclusion criteria required participants to have primary responsibility for their child’s health care decisions.

**Table 1.** Sociodemographic characteristics of participants.

| Participant ID | FGD ID | Gender | Age range | Number of Children |
| --- | --- | --- | --- | --- |
| 001 | B1 | Female | 31-35 | 9 |
| 002 | B1 | Female | 26-30 | 5 |
| 003 | B1 | Female | 31-35 | 7 |
| 004 | B1 | Female | 41-45 | 9 |
| 005 | B1 | Female | 41-45 | 8 |
| 006 | B1 | Female | 41-45 | 4 |
| 007 | B1 | Female | 21-25 | 8 |
| 008 | B1 | Female | 26-30 | 2 |
| 009 | B2 | Male | 41-45 | 8 |
| 010 | B2 | Male | 36-40 | 6 |
| 011 | B2 | Male | 26-30 | 2 |
| 012 | B2 | Male | 26-30 | 3 |
| 013 | B2 | Male | 21-25 | 4 |
| 014 | B2 | Male | 26-30 | 3 |
| 015 | B2 | Male | 31-35 | 8 |
| 016 | B2 | Male | 21-25 | 5 |
| 017 | B3 | Female | 26-30 | 4 |
| 018 | B3 | Female | 46-50 | 10 |
| 019 | B3 | Female | 31-35 | 3 |
| 020 | B3 | Female | 41-45 | 6 |
| 021 | B3 | Female | 21-25 | 1 |
| 022 | B3 | Female | 18-20 | 2 |
| 023 | B3 | Female | 46-50 | 9 |
| 024 | B3 | Female | 51-55 | 10 |
| 025 | Y1 | Female | 21-25 | 3 |
| 026 | Y1 | Female | 36-40 | 5 |
| 027 | Y1 | Female | 21-25 | 2 |
| 028 | Y1 | Female | 21-25 | 1 |
| 029 | Y1 | Female | 21-25 | 1 |
| 030 | Y1 | Female | 21-25 | 1 |
| 031 | Y1 | Female | 21-25 | 3 |
| 032 | Y1 | Female | 21-25 | 1 |
| 033 | Y2 | Female | 41-45 | 6 |
| 034 | Y2 | Female | 21-25 | 3 |
| 035 | Y2 | Female | 21-25 | 2 |
| 036 | Y2 | Female | 36-40 | 6 |
| 037 | Y2 | Female | 36-40 | 3 |
| 038 | Y2 | Female | 36-40 | 4 |
| 039 | Y2 | Female | 26-30 | 3 |
| 040 | Y2 | Female | 21-25 | 2 |
| 041 | Y3 | Male | 31-35 | 7 |
| 042 | Y3 | Male | 31-35 | 4 |
| 043 | Y3 | Male | 36-40 | 5 |
| 044 | Y3 | Male | 36-40 | 8 |
| 045 | Y3 | Male | 41-45 | 17 |
| 046 | Y3 | Male | 46-50 | 18 |
| 047 | Y3 | Male | 36-40 | 11 |
| 048 | Y3 | Male | 36-40 | 7 |
| 049 | A1 | Female | 41-45 | 7 |
| 050 | A1 | Female | 31-35 | 6 |
| 051 | A1 | Female | 31-35 | 2 |
| 052 | A1 | Female | 36-40 | 5 |
| 053 | A1 | Female | 26-30 | 2 |
| 054 | A1 | Female | 18-20 | 2 |
| 055 | A1 | Female | 18-20 | 2 |
| 056 | A1 | Female | 21-25 | 2 |
| 057 | A2 | Female | 31-35 | 7 |
| 058 | A2 | Female | 46-50 | 3 |
| 059 | A2 | Female | 46-50 | 5 |
| 060 | A2 | Female | 36-40 | 5 |
| 061 | A2 | Female | 41-45 | 3 |
| 062 | A2 | Female | 26-30 | 1 |
| 063 | A2 | Female | 46-50 | 3 |
| 064 | A2 | Female | 46-50 | 7 |
| 065 | A3 | Male | 21-25 | 1 |
| 066 | A3 | Male | 31-35 | 2 |
| 067 | A3 | Male | 26-30 | 2 |
| 068 | A3 | Male | 61-65 | 11 |
| 069 | A3 | Male | 36-40 | 3 |
| 070 | A3 | Male | 51-55 | 11 |
| 071 | A3 | Male | 31-35 | 3 |
| 072 | A3 | Male | 26-30 | 1 |

### Data collection

Data were collected through nine focus group discussions (FGDs) with caregivers from three different communities. Focus groups were used to encourage discussion and exchange of ideas among participants, and to promote a deeper understanding of shared experiences and perspectives. Each FGD included 8 participants and lasted approximately 60-90 minutes. Discussions were facilitated by trained researchers using a semi-structured interview guide. The FGDs were conducted in local languages and audio recorded for transcription. Field notes were also taken during the discussions to capture non-verbal cues and contextual observations. FGDs with women were conducted by a female moderator and FGDs with men were conducted with a male moderator.

### Data analysis

A content thematic analysis approach was used to analyse the qualitative data. Audio recordings were transcribed verbatim from Hausa and translated into English. The transcripts were checked for accuracy by bilingual researchers. The content analysis followed the steps outlined by Clarke & Braun (2017). Two independent researchers (co-authors XX. XX) analysed the data by reading and rereading the transcripts and field notes. Key phrases and statements were systematically coded to capture meaningful segments related to the research questions. Themes were refined and defined to ensure that they clearly encapsulated the findings. Final data analysis was agreed by consensus resulting from iterative discussions between co-authors (xx, xx, and xx). Data analysis was conducted using qualitative analysis software (NVivo).

### Ethics approval and consent to participate

Ethical approval for this study was obtained from the Ethical Committee from the University of xx (IRB statement xx) and the National Health Research Ethics Committee of Nigeria (xx).

### Consent for publication and participation

Written informed consent was obtained from all participants prior to data collection. Confidentiality was maintained by anonymising participant data, and participants were informed of their right to withdraw at any time without any consequence. As the manuscript includes individual-level qualitative data (e.g. direct quotations), consent for publication was also obtained from all participants. The study adhered to the Consolidated Criteria for Reporting Qualitative Research (COREQ) (60) checklist to ensure transparency and rigour in reporting the methodology and findings. This included documenting details of the research team, data collection and analysis processes, and providing a clear audit trail of coding and theme development.

### Reflexivity statement

Reflexivity was an essential component throughout our research process, guiding our methodological choices, data analysis and interpretation of findings. Our research team included Nigerian and international researchers with expertise in global health and qualitative research methods; the Nigerian team was also fluent in the languages and familiar with the cultural and social context of northeast Nigeria. Throughout data collection the Nigerian researchers (co-authors xx, xx, xx, xx, xx, xx) took care to minimize potential power dynamics by ensuring that focus group facilitators were culturally and linguistically matched to participants. Regular team meetings were held to critically reflect on data collection experiences, field notes and emerging themes, helping us to remain aware of our own potential biases and assumptions.

## Results

A total of nine focus group discussions (FGDs) were conducted in June 2024 across three states in northeast Nigeria: three FGDs each in Borno, Yobe, and Adamawa. Six FGDs were composed exclusively of women (n=48), while three FGDs included only men (n=24), resulting in a total of 72 participants (see Table 1). Participants reported having between 1 and 18 children, with a total of 355 children across all participants. Although most participants reported having an occupation, women were more likely than men to report no formal occupation or any means of livelihood. Participants ranged in age from 20 to 61 years, with an average age of 34 years.

The analysis of the nine focus group discussions (FGDs) with caregivers of children under five in conflict-affected regions of northeast Nigeria revealed a complex interplay between health-seeking behaviour, trust in health systems and the impact of ongoing violence. Participants highlighted significant barriers to accessing health care, enabling strategies developed within their communities, and mixed attitudes towards vaccination. In addition, the role of health information dissemination and its influence on health decisions emerged as a key factor. The findings are organised into four major themes (Table 2, and Figure 1): i) barriers to accessing healthcare, ii) community resilience and adaptation, iii) attitudes towards vaccination, and iv) the health information landscape.

**Figure 1.**
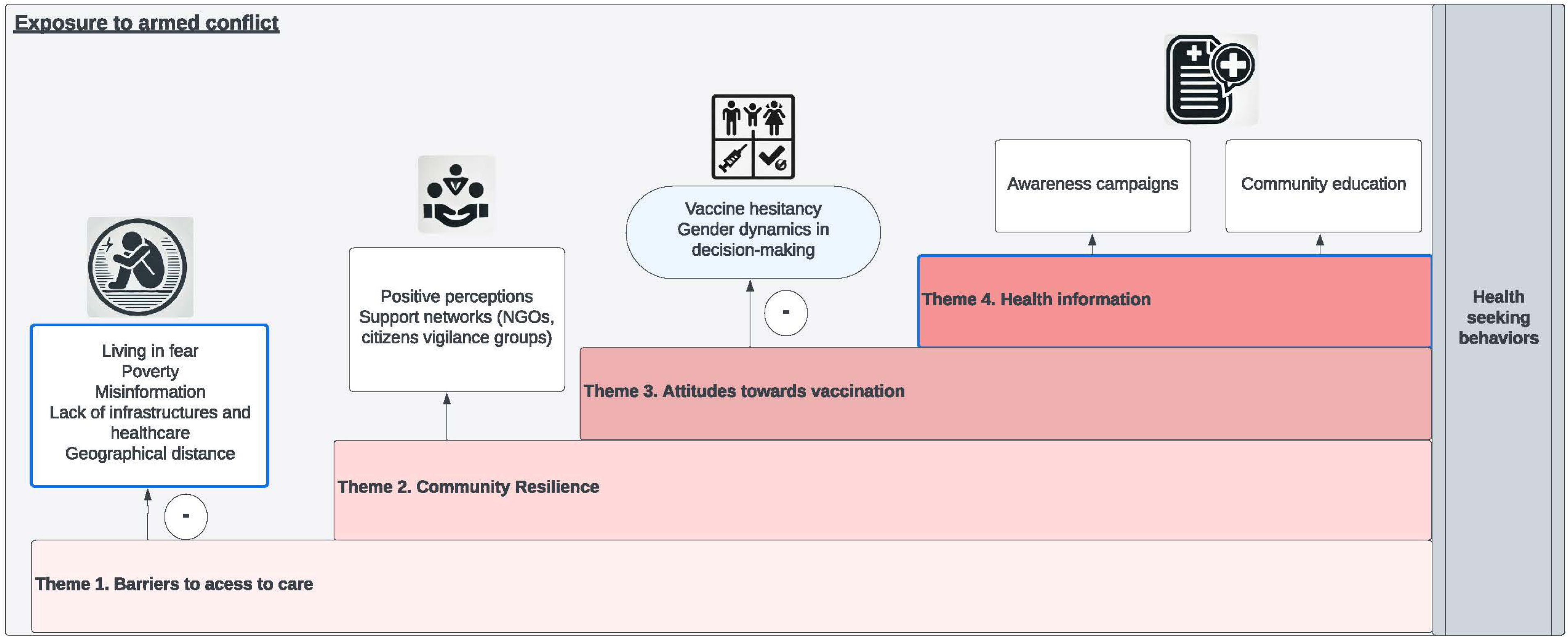
Main themes of health seeking behaviour of caregivers with children under 5 years old.

**Table 2.** Description of themes.

| THEMES | Description |
| --- | --- |
| **1. Navigating barriers to accessing healthcare** | This theme describes main barriers of access to care. Caregivers consistently identified multiple barriers to accessing health services, exacerbated by the ongoing conflict. Fear of violence and insecurity often obstructed caregivers from visiting health facilities, further complicated by the destruction of infrastructure and the absence of health workers. Economic challenges, including widespread poverty and the cost of transport and treatment, were significant barriers. In addition, long distances to health facilities posed logistical and safety concerns, especially in emergencies. Together, these barriers limited caregivers' ability to seek timely and appropriate care. |
| *1.1 Living in fear and insecurity* |  |
| *1.2 Economic constraints* |  |
| *1.3 Infrastructural deficits* |  |
| *1.4 Geographical distance* |  |
| **2. Resilience and community enabling strategies** | This theme describes collective mechanisms of resilience during conflict times. In the midst of the inherent conflict-related challenges, caregivers showed remarkable resilience through community-based strategies. Participants described how vigilante groups provided security and facilitated access to health care by escorting patients. Communities pooled resources for transportation and medicines, while local NGOs filled critical gaps left by the government, providing essential health services and support. Where formal healthcare was inaccessible, traditional herbalists were used as alternative. These enabling measures underscored the resourcefulness of communities in overcoming systemic failures in health care. |
| *2.1 Experiences of violence* |  |
| *2.1 Support networks* |  |
| *2.3 NGO involvement* |  |
| *2.4 Integration of traditional practices* |  |
| **3. Attitudes towards vaccination** | This theme describes attitudes regarding vaccination. Caregivers were generally positive about it, recognising its importance in preventing disease and protecting children's health. However, significant barriers remained, including disruption of immunisation services due to conflict, and vaccine hesitancy due to fear of adverse effects. Gender dynamics influenced decision-making, with male household members often having the authority to agree or refuse vaccination. Community-led awareness campaigns played a key role in addressing these concerns, increasing acceptance of the vaccine and improving uptake rates. |
| *3.1 Positive perceptions* |  |
| *3.2 Vaccine hesitancy* |  |
| *3.3 Gender dynamics in decision-making* |  |
| **4. Health information landscape** | This theme addresses health information. Participants highlighted the effectiveness of awareness-raising campaigns, particularly those involving trusted community leaders or broadcast on radio, in increasing knowledge about maternal and child health. These efforts helped to counter misinformation, particularly around immunization, but gaps in access to reliable information remained. Participants stressed the importance of sustained, context-specific health education initiatives to further improve health decision-making and outcomes in their communities. |
| *4.1. Role of awareness campaigns* |  |
| *4.2 Community education* |  |

### Theme 1: Navigating barriers to accessing healthcare

This theme describes main barriers of access to care (see Table 3 for further citations).

**Table 3.** Barriers to healthcare (Theme 1)

|  |
| --- |
| **1 Navigating barriers to accessing healthcare** |
| **1.1 Living in fear and insecurity** |
| [1.1a] "In this situation, the armed conflict, banditry and thieves really affected us seriously and it made it very difficult for us to access the healthcare services, because of this situation, even if you are sick in the night no one or not even an Okada (motorcycle) will agree to take you to the hospital because of fear and lack of trust on who is picking so it really affected our healthcare access in this community." FGD2, P1<br>[1.1b] "There was a time we were at home around 7.00pm we heard gunshots and were told to run away but I could not run away I stayed on top of a hill outside the community. That time I was nine months pregnant, and because of that panic I delivered in the bush inside a trench." FGD3, P8. |
| **1.2 Economic constraints** |
| [1.2a] "When the insurgency started in our area, I was on the farm, I ran away and we didn't go back home. At that time, we didn't have money, my husband only had N300 naira in his pocket so we managed to go to his village where he left me under the care of his caregivers and went to Lagos. So, after he came back from Lagos we returned back to our village and after some time, these insurgents attacked us again and in the event my husband was killed by the Boko Haram insurgents." FGD3, P1.<br>[1.2b] "We lived in difficulties during the violent conflict and to access healthcare was very difficult because both my husband and my caregivers had fled. Yes, our health seeking behaviour has changed because we have not been able to access free healthcare services, we are even afraid to go out and take ourselves and our children to the hospital." FGD7, P8<br>[1.2c] "One of the barriers is poverty that stop caregivers from taking their children for vaccination; lack of involvement of community leaders is another factor; distance to the hospital is also another reason; and lack of transport can be a factor. Lack of provision of free drugs is another reason." FGD8, P2<br>[1.2d] "Poverty and financial constraints are the main barriers that prevent caregivers from taking their children for vaccination." FGD8, P3<br>[1.2e] "Some would like to visit hospital but lack of money prevents them from accessing care." FGD1, P3 |
| **1.3 Infrastructural deficits** |
| [1.3a] “There was a time when my child had an accident and was taken to the health facility but there were not enough working materials to treat him. Then we were referred to secondary and then to tertiary institutions to treat him, so my health seeking behaviour has not changed during the violent conflict.” FGD1, P3<br>[1.3b] “At that time, we went back there was no access to healthcare services because the health facilities were not operating and at the same time there was no potable drinking water, and this seriously affected our healthcare services. There was no food, and everything was in shambles there was no access to portable water, electricity and no good healthcare service.” FGD3, P5 |
| **1.4 Geographical barriers** |
| [1.4a] “Yes, at that time there was no means of transport to take your child or pregnant woman to the hospital at that time. We really lived in a difficult situation because my husband has run away to Enugu, and I have stopped hearing from him since the armed conflict began. I am now pregnant, and my expected date of delivery (EDD) is this month of May (2024), I don’t know what to do, I am living in confusion. But at times my mother-in-law is assisting me with the little she gets.” FGD1, P5.<br>[1.4b] “Yes, for those that live in villages under this community it is not easy for them to transport themselves to the health facility because their houses are very from the hospital is one of the barriers they are facing. ” FGD 1, P3<br>[1.4c] “Yes, our health seeking behaviour has changed due to the fear of Boko Haram and also there was problem of transportation because of distance and also some other people live in the hard-to-reach areas. That is why our seeking behaviour has changed and some could not access care in the hospital.” FGD6, P8 |

### 1.1 Living in fear and insecurity

Participants described the overwhelming fear and insecurity caused by the ongoing violence, which significantly limited their ability to seek health care. The threat of being attacked, kidnapped or mistaken for insurgents created an environment of constant vigilance, making visits to health facilities fraught with fear and danger. For many caregivers, fear extended beyond their personal safety to concerns about the well-being of their children in their absence from home:

> “We were afraid of been attacked on our way to the hospital and at the same time we were afraid that our children could be kidnapped by the bandits in our absence.” FGD1, P8

The danger was worst at night, as caregivers completely lacked transportation options [1.1a]. In some cases, the insecurity disrupted maternal healthcare entirely. One participant shared a traumatic experience of being forced to deliver her child in the woods while running away from an armed attack [1.1b].

### 2 Economic constraints

Economic hardship emerged as another barrier to healthcare access, with poverty often cited as a primary factor limiting the ability of caregivers to seek necessary treatment. Namely how poverty affected their ability to afford transportation, medical bills or medication, leaving many without access to essential health services. One participant described how the loss of livelihoods and personal tragedies aggravated their struggles [1.2a]. The destruction caused by the conflict left many participants even more impoverished and unable to meet ends:

> “*The armed conflict really affected us seriously and it was difficult for us to access any form of healthcare services, especially anything that is not an emergency. We live in poverty because they have burnt everything, we are left with nothing; even to buy medicine we can’t afford*.” FGD7, P3

Financial constraints not only affected access to care, but also disrupted health-seeking behaviour [1.2b] and access to basic healthcare services, such as routine vaccinations for children [1.2c]. The cumulative effect of financial hardship was briefly summarized by several participants [1.2d], [1.2e].

### 3 Infrastructural deficits

The armed conflict severely affected the accessibility and functionality of health facilities. Participants described how the destruction of infrastructure, adding to the migration of health workers and shortages of medical supplies created significant barriers to accessing timely and appropriate care, particularly during critical emergencies:

> *“Another issue that we were exposed during the violence was the lack of enough healthcare providers at the hospital both during the day and at night; most of them have either migrated or fled to other places for their safety.* “ FGD1, P1

Inadequate resources in health facilities forced caregivers to navigate complex referral systems, often without resolution, as described in one case of emergency care for a child [Table 3, 1.3a]. Participants also detailed how the lack of basic amenities such as clean water, electricity, and operational health facilities exacerbated the crisis [Table 3, 1.3b]. The combination of inadequate medical supplies and staff, coupled with wider socio-economic hardship, had devastating consequences, including deaths, particularly for children with malaria and typhoid, as tragically reported by participants:

> *“But on our return to our community, access to healthcare became very difficult. This insecurity issues and violent conflict has affected us seriously. At that time, my child was affected by malaria and typhoid but due to lack of care from the hospital and the inability of healthcare, he died.” FGD3, P3*

### 4 Geographical barriers

Long physical distance to healthcare facilities was another significant challenge. One participant highlighted the isolation experienced during pregnancy due to lack of transport and family support, exacerbated by displacement [1.4a]. For those living in villages far from healthcare facilities, the distance itself became a barrier to accessing care [1.4b]. Participants emphasized how poor road conditions, particularly during the rainy season, limited access to healthcare. The combination of fear to dislocate and geographic isolation on those living in hard-to-reach areas further discouraged caregivers from seeking care [1.4c].

### Theme 2: Resilience and community enabling strategies

This theme describes experiences of violence and consequential coping mechanisms of resilience during conflict times (see Table 4 for further citations).

**Table 4.** Community Resilience (Theme 2)

|  |
| --- |
| **2. Resilience and community enabling strategies** |
| **2.1 Experiences of violence** |
| [2.1a] “Exposure to violence has affected our well-being in the sense that after my husband was killed by the Boko Haram insurgents, I am left to take care of my children, their schools fees, shelter, health and any other well- being. This is very difficult for me and I encountered serious challenges been a woman without a husband with some kids under my care. Yes, there are few people in the community that support me but not 100%.” FGD1, P8<br>[2.1b] “There were killings of innocent people within the community after we returned around April this year (2024). I witnessed and saw them lying on the ground dead; about 20 people were killed (both males and females including young children) within or in some parts of our community after we have returned. As a result of that tragedy, accessing healthcare services become difficult because of fear and anxiety.” FGD3, P1.<br>[2.1c] “Yes, I was exposed to the violent conflict of Boko Haram, my husband ran away and left us with our children, and some were even shot, and my husband was kidnapped and stayed with them for over one year, he was later released.” FGD4, P2.<br>[2.1d] “When I was in Gwoza, I was exposed to Boko Haram insurgency where my father was killed by the insurgents and my husband was also killed. It was an extremely terrifying period.” FGD4, P3. |
| **2.2 Support networks** |
| [2.2a] “Yes, there is committee of hunters and vigilantes that patrols the area in the night so that those bandits do not attack our community. They also have the support of government security agencies in protecting the community.” FGD2, P6<br>[2.2b] “We used a “No Boko Group” that assists in protecting the community and in fighting with the armed bandits. We also pay them some allowance at the end of every month and salary/allowance come through the emir’s palace, stakeholders, and philanthropists and this will motivate them to protect the community.” FGD2, P7<br>[2.2c] “We built a gate at the entrance of the community so that any person that is coming into the community will be monitored and security is manned at the gate entrance and this has improved the security situation of our community.” FGD2, P8 |
| [2.2c] “There is a group of youths that used to escort us also in the night if there is a person that is seriously sick or when a woman is in labour.” FGD9, P4 |
| **2.3 NGO involvement** |
| [2.3a] “At that time my daughter was sick we were unable to access care in the hospital, but we have the numbers of their phone but when the illness became complicated, we were asked to go to Maiduguri for further care. We went to one NGOs hospital in Maiduguri, and we received good medical care at that time.” FGD4, P4<br>[2.3b] “I think these NGOs are better than the government but also the government is trying.” FGD4, P1<br>[2.3c] “But on our return to our community, accessing healthcare became very difficult. There were no adequate drugs in the health facility we attend, no adequate healthcare personnel. We don't even have money to buy the medicines, let alone to transport ourselves to the hospital. This insecurity issues and violent conflict have affected us seriously. At that time my child was affected by malaria and typhoid, but due to lack of care from the hospital and the inability of professional health personnel, he died.” FGD3, P3 |
| **3.4 Integration of traditional practices** |
| [3.4a] “What made us change our health-seeking behaviour was that if you went to the hospital, you would not see any health professional on duty. So, we go back to the herbalist woman to check our pregnancy.. “ FGD3, P4 |

### 1 Experiences of violence

Caregivers shared devastating accounts of violence and personal loss, illustrating how exposure to the conflict has profoundly affected their ability to care for their families and access health care. Many participants described how the violent insurgency had disrupted their lives, leaving them to cope with the loss of loved ones, emotional trauma and significant socio-economic challenges. For some, the loss of a spouse placed an immense burden on them as the sole caregiver, adding to the difficulties of managing daily life [2.1a]. The psychological and practical toll of losing family members to insurgent attacks was often mentioned:

> “*I felt bad because I have orphans that I am now taking care of, which is very difficult for me. My husband was a state security personnel, they came and shot him together with his friend and I can say it was a bad moment for me.*” FGD1, P4

Participants also described how witnessing mass violence and killings within their communities caused fear and anxiety [Table 3, 2.1b]. The trauma of abduction, displacement, and family separation due to the insurgency added layers of hardship to access care [Table 3, 2.1c], [Table 3, 2.1d].

### 2 Support networks

The destructive impact of violence on caregivers’ lives, affected their physical and mental health and compounded barriers to healthcare access. Despite these challenges, many caregivers displayed resilience, enabling continuity of care for their families. Caregivers described how their communities had organised themselves, forming vigilante groups and other initiatives to address security concerns and assist those in need. These networks provided practical support, including transportation, financial assistance and even protection during medical emergencies. Vigilant groups played a central role in improving security and access to health care [Table 3, 2.2a]:

> “*We formed a group of vigilantes that control the security situation in the night and when the bandits come, they would engage in a fight with them, so that how we can have peace in the community against the attack of armed bandits. As a result, many neighbouring communities did the same*.” FGD2, P4

The "No Boko Group," a community-formed security initiative, was specifically praised for its contributions to safety and healthcare access, including its financial sustainability model [Table 3, 2.2b]. Physical security measures, such as building a monitored community gate, also contributed to a safer environment for residents [Table 3, 2.2c]. For medical emergencies, youth groups provided critical escort services, ensuring safe transportation for patients and expectant mothers [Table 3, 2.2c]. These support networks highlight the resilience and resourcefulness of communities in mitigating the impact of conflict. By organizing local initiatives and leveraging collective resources, these communities were able to address some of the barriers to healthcare access and create a sense of security.

### 3 NGO involvement

Non-governmental organisations (NGOs) emerged as key actors in addressing the health needs of conflict-affected communities, stepping in where government services were absent or inadequate. In several cases, NGOs provided critical care when local facilities were inaccessible or overwhelmed. One participant described how an NGO hospital in Maiduguri became a lifeline when her daughter’s illness required more advanced medical care [Table 3, 2.3a]. For others, NGOs played a crucial role in accessing the health care during displacement:

> “*At that time when I was sick, there were doctors that used to treat us in the community. My child also suffered from malaria when we slept in the bush and we were all taken to the hospital by the NGOs*.” FGD4, P4

Participants contrasted the reliability of NGOs with government efforts, noting that while the government made some attempts to provide services, NGOs often outperformed them in both delivery and responsiveness [Table 3, 2.3b]. Therefore, participants described the vital role of NGOs in bridging gaps left by government inaction, particularly during times of acute crisis.

### 4 Integration of traditional practices

Whether because of the absence of accessible formal health services or cultural practice, caregivers often turned to herbalists and traditional medicine to address urgent health needs. Participants highlighted how the lack of available healthcare providers and the disruption of formal medical services during the conflict led many to rely on these traditional practices:

> “*It doesn’t affect me but it affected some of the neighbours and they could not seek care at that time, so their health seeking behaviour changed because they resort to visiting herbalist for care at that time*.” FGD6, P6

Another participant described how the unavailability of healthcare providers at local hospitals forced a change in health-seeking behaviour, resulting in the use of traditional methods for essential care such as pregnancy monitoring [Table 3, 3.4a]. Participants emphasized that access to formal healthcare was not only a necessity but also a marker of peace and stability in their communities.

### Theme 3: Attitudes towards vaccination

This theme describes attitudes regarding vaccination (see Table 5 for further citations).

**Table 5.** Attitudes towards vaccination (Theme 3)

|  |
| --- |
| **3. Attitudes towards vaccination (Theme 3)** |
| **3.1 Positive perceptions** |
| [3.1a] “The majority of people now understand the importance and benefits of vaccination for their children. In the past people did not believe in vaccines due to ignorance, illiteracy, and lack of awareness about the benefits of vaccines. In the past, people were hiding their children from vaccination. But when the polio vaccine started having a positive effect on our children’s suffering from polio, other caregivers came to understand about the importance of vaccines, and they started allowing their children to be vaccinated. People thought that the government was trying to reduce the number of the people in a society, but due to awareness campaigns they now understand that this is not true. As for the risk, I don’t think vaccination has any risk to the lives of our children. The only risk I can say is that if the doctor or healthcare worker is not an expert in vaccination, he might do it in the wrong way or place and this will make a child’s leg to swell”. FGD2, P8<br>[3.1b] “I am a teacher and at times *vaccinators* used to visit our schools to vaccinate our children against polio and other related diseases. Recently, as I recall, there was an intervention campaign on vaccines for 9 and 14 year-olds and no one agreed to be vaccinated. And the headmaster did not allow them to vaccinate the children because he was having challenges from the caregivers of the children. The vaccine is to prevent cervical cancer, but some agreed to be vaccinated and others refused. The reason for their refusal is also a result of rumours about the side effects of the vaccine.” FGD1, P4 |
| **3.2 Misinformation as a barrier** |
| [3.2a] “Usually what makes them not to vaccinate their children is the adverse effect of some vaccines like DPT, which makes the child cry or fall sick and have a swollen leg. This most of times stops them from coming back to take or complete the vaccination.” FGD3, P3<br>[3.2b] “Some are suspecting that the vaccine is used for family planning so that in the future their children would not bear children again.” FGD4, P8<br>[3.2c] “There is a lack of awareness about the importance of the vaccine. This is the reason why some do not bring their children for vaccination. It is ignorance and illiteracy.” FGD5, P2<br>[3.2d] “The lack of free nutritional supplements for the children is what discourages caregivers from taking their children for vaccination. Because in the past children were given incentives after each dose of vaccine.” FGD8, P2<br>[3.2e] “Yes, there is believe that it is plot to decrease the population of our people in the community. That is why some caregivers don’t take their children for vaccination.” FGD8, P1 |
| [3.2f] “The reason why some people do not like to vaccinate their children as I have said is illiteracy, ignorance and lack of awareness about the importance and benefits of vaccination. Everybody knows prevention is better than cure, so for those who do not allow their children to be vaccinated, it is mainly due to illiteracy.” FGD2, P7<br>[3.2g] “But in my own view, this issue of exclusive breastfeeding is the one that has made our children to misbehave and do not respect the elders because they were not given water when they were born as the western people directed. That is the reason why now most of the youths whose mothers practiced exclusive breastfeeding do not respect their elders.” FGD1, P4<br>[3.2h] “Some of their husbands do not allow them to vaccinate their children and they think that it is going to make their children sterile in the future.” FGD4, P4 |
| **3.3 Gender dynamics in decision-making** |
| [3.3a] “Yes, the decision to seek health might change because of lack of transport to take your wife to the hospital for maternal healthcare; but for me, nothing has changed because I must find a way of taking my family to the hospital to access care services, be it maternal or any other healthcare services that we need. Yes, I am afraid but I had no choice but to seek healthcare service(s) at that particular time.” FGD2, P8<br>[3.3b] “In fact, even the husbands that do not allow their wives to take their children for immunisation now all agree and even encourage it. There is a lot of awareness.” FGD5, P2<br>[3.3c] “One of the reasons is that my husband doesn’t like it because of the side effects. But my in-laws make sure that our children are vaccinated.” FGD9, P3<br>[3.3d] “My in-laws do not like our child to be vaccinated, but I refused their advice and took my child for vaccination.” FGD9, P4<br>[3.3e] “There is need for increased awareness in the community about the importance and benefits of vaccines, and the majority of caregivers who have doubts about vaccines are fathers or husbands. In my view, their concern is related to tradition and religious beliefs that prevent them from allowing their children to be vaccinated. But there is a commitment from the chief Imams of the area to include the issue of vaccination in their sermons, telling the public about the importance of the vaccine for people.” FGD1, P6 |

### 1 Positive perceptions

Participants frequently acknowledged the critical role of vaccinations in improving child health and eradicating diseases. Awareness campaigns and community engagement were cited as key factors in shifting perceptions and increasing acceptance of vaccines. One participant noted the transformative impact of vaccination campaigns and the involvement of community leaders:

> “*What I know about the importance of vaccination for our children, is that it is used to prevent polio, measles, meningitis. The health of our children has improved because most of those diseases have been eradicated because of vaccination. This was a result of raising awareness in the community either in the mosque, churches and social gatherings*.” FGD2, P5

The historical barriers to vaccination, such as ignorance and misinformation, and how these have been mitigated through education efforts was also mentioned [Table 5, 3.1a]. Notably, a new term *vaccinators* emerged as a local expression used by some participants to refer to healthcare workers who go door to door for vaccination [Table 5, 3.1b].

### 2 Misinformation as a barrier

Participants highlighted a number of misconceptions and beliefs that fuelled vaccine hesitancy, including suspicions about the intentions behind vaccination programmes, and a lack of awareness of their benefits. Some participants highlighted the role of rumours circulating about the safety and efficacy of vaccines, which led to mistrust among caregivers:

> “*Their reasons are that they believe the rumours circulating about the side effects and efficacy of vaccines. That is why they do not allow their children to be vaccinated*.” FGD1, P3

Fear of side effects, particularly after receiving certain vaccines, also discouraged caregivers from continuing or completing vaccination schedules [Table 5, 3.2a]. Deep-rooted suspicions or myths about the purpose of vaccines added to the hesitancy, with some caregivers believing that vaccination was linked to family planning or efforts to reduce population growth [Table 5,

3.2b]. Participants also pointed to broader issues of illiteracy and lack of awareness as key drivers of vaccine hesitancy [Table 5, 3.2c]. Finally, participants noted that the withdrawal of incentives during conflict previously provided alongside vaccination - such as nutritional supplements - discouraged some caregivers from participating in immunization programs [Table 5, 3.2d, 3.2e, 3.2f, 3.2g, 3.2h].

### 3. Gender dynamics

Cultural norms and traditional gender roles meant that women’s access to health care and vaccination services often depended on the consent of their husbands or in-laws. Participants shared mixed experiences of male involvement in health care decisions during conflict. While some male caregivers expressed their commitment to ensuring that their families had access to health care despite challenges, others restricted their wives’ or children’s access due to fears or traditional beliefs [Table 5, 3.3a]. In contrast, other participants revealed how concerns over safety and cultural beliefs restricted women’s healthcare access, even for necessary services [3.3b]. Women themselves sometimes resisted family pressures and ensured their children received vaccinations despite opposition from husbands or in-laws [Table 5, 3.3c, 3.3d, 3.3e].

“*For me, I will not allow my wife to attend ANC services during these times of conflict anyway, but I prefer to visit the hospital during daylight because of the insecurity. Yes, they attend ANC but there is fear; but there was nothing they could do.*” FGD2, P8

### Theme 4: Health information landscape

Dissemination of health information emerged as a critical factor in shaping health-seeking behaviour (see Table 6 for further quotes).

**Table 6.**
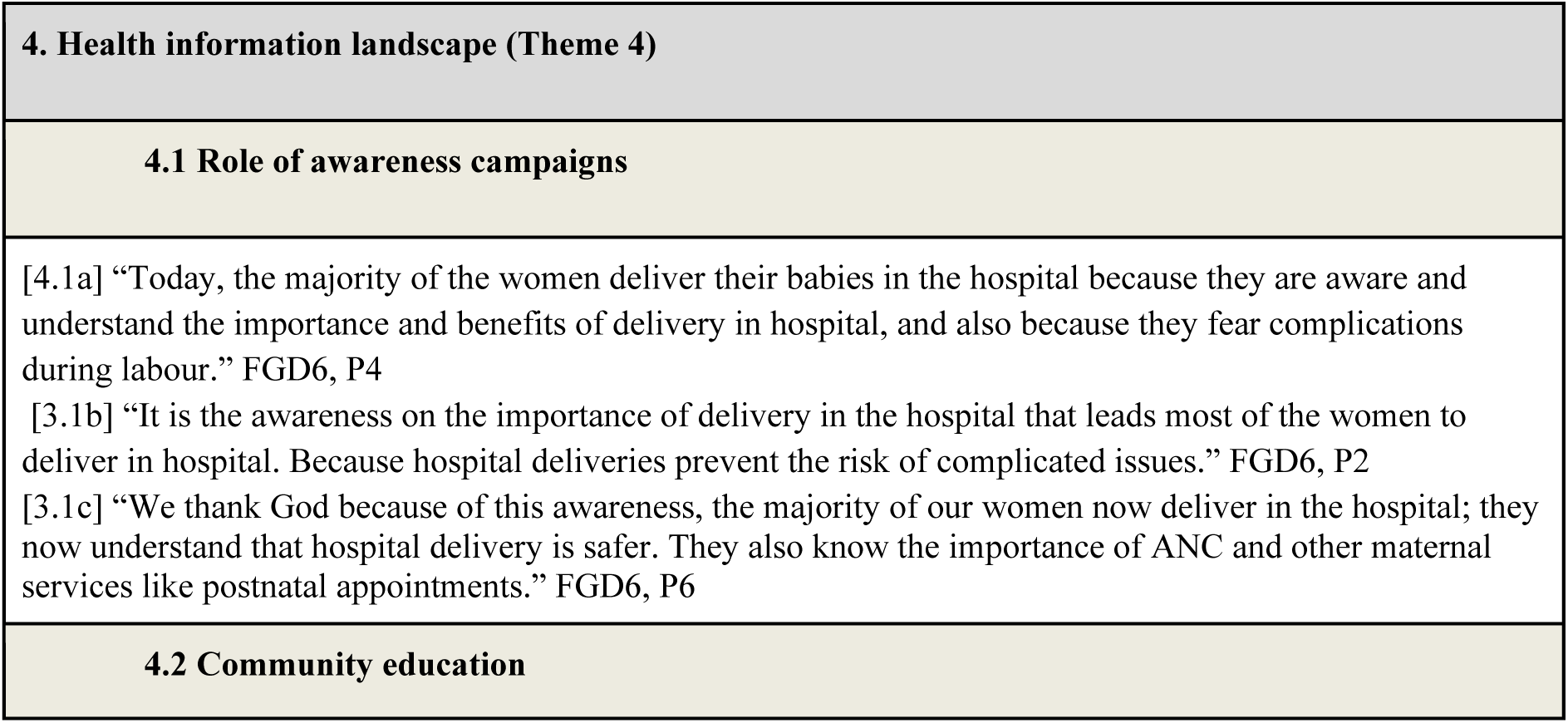
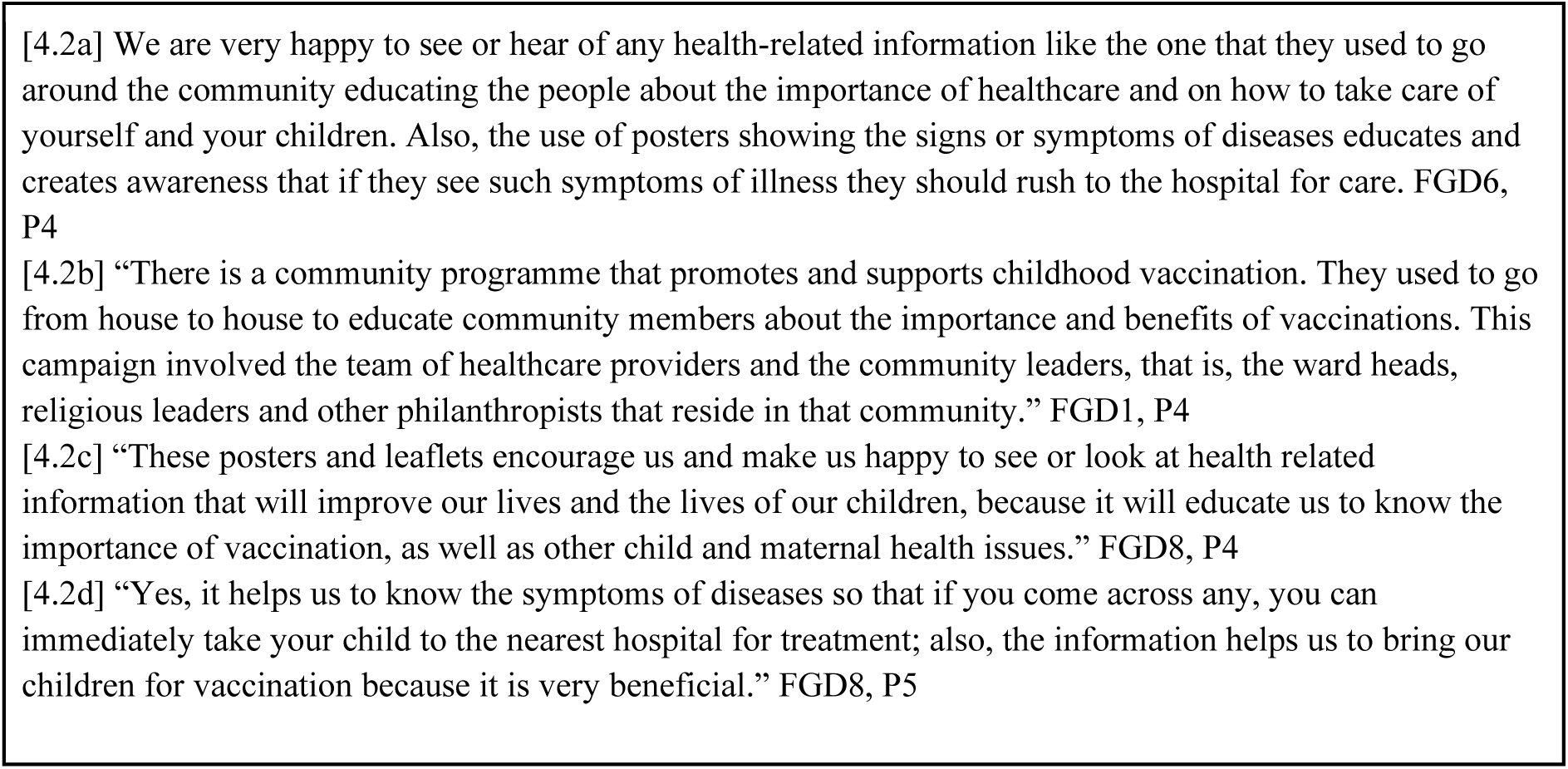
Health information landscape (Theme 4)

### 1 Role of awareness campaigns

Participants highlighted the positive impact of health promotion efforts, particularly those involving trusted community leaders and delivered through accessible channels such as radio [Table 6, 4.1a]. Participants attributed the shift towards hospital deliveries directly to these targeted health promotion campaigns, which emphasised the safety of institutional delivery [Table 6, 4.1b]. In addition to increasing hospital deliveries, awareness campaigns were recognized for promoting other maternal health services, such as antenatal and postnatal care [Table 6, 4.1c].

### 2 Community education

Specifically, it was mentioned the importance of door-to-door campaigns, visual aids such as posters, and the involvement of local leaders in providing health information. One participant expressed appreciation for education efforts that emphasised early recognition of disease symptoms and the importance of accessing health services [Table 6, 4.2a]. House-to-house campaigns were described as effective, involving healthcare providers, community leaders and local philanthropists to build trust and promote vaccination uptake [Table 6, 4.2b]. Participants also liked visual materials like posters and pamphlets for their ability to communicate health messages effectively and encourage maternal and child health practices [Table 6, 4.2c, 4.2d].

## Discussion

This study has provided a comprehensive overview of health-seeking behaviour, trust in health systems and vaccine hesitancy among caregivers of children under five in conflict-affected regions of northeast Nigeria. Our findings showed the multiple challenges caregivers faced in accessing health care amidst ongoing violence, poverty, and systemic mistrust of institutions. Very importantly, pervasive insecurity, characterised by the daily threat of attack and kidnapping, has created a climate of fear that limited mobility and disrupted access to healthcare. Consistent with findings from other conflict-affected regions (17), caregivers reported that these fears often led to delays or avoidance of needed healthcare, particularly maternal and child health services. Economic constraints compounded these challenges, with caregivers often unable to afford transportation, medical fees, or medication, as also observed in other studies (42). Despite these challenges, caregivers showed remarkable resilience, using community-driven strategies to mitigate the impact of conflict on access to health care. The formation of vigilant groups and community security initiatives, such as the ’No Boko’ groups, exemplifies the ability of local communities to address security concerns and facilitate access to health services. These findings are consistent with the literature highlighting the importance of local resilience in conflict settings (43). Non-governmental organisations (NGOs) also played a key role, filling critical gaps left by the government and providing essential health services, particularly during emergencies. However, trust in health systems emerged as a major key issue influencing caregivers’ health decisions and vaccine uptake. Erosion of trust due to conflict-related disruptions, misinformation and historical events such as the 2003 polio vaccine boycott (44) contributed to vaccine hesitancy. While caregivers generally recognised the benefits of vaccination, fear of side effects, suspicions about vaccine safety and patriarchal decision-making structures often led to hesitation. Community-based awareness campaigns and the involvement of religious and community leaders were instrumental in rebuilding trust and promoting vaccination, as noted in other settings (45). The dissemination of accurate and accessible health information was also identified as a key enabler of positive health-seeking behaviour. Similarly, campaigns led by trusted community figures and tailored to local contexts effectively countered misinformation and improved maternal and child health practices (46). Finally, our research showed that community resilience resulted not only from physical infrastructures, but also from social capital and enabling informal solutions. In particular, bidirectional capacity building was found to be critical in acute crises, fostering mutual trust within the community. This finding is consistent with broader research on community engagement in health system interventions in conflict-affected regions. A scoping review comprising 19 studies addressing community engagement in access to care in conflict-affected areas (47), included one study focused on health system resilience in the same setting as our study (Yobe State) (48). Their findings resonated strongly with ours, namely, how community-organised transport networks emerged to overcome mobility restrictions imposed by the insurgency, and also, how bottom-up collaboration can strengthen crisis response mechanisms while maintaining access to care.

Adding to previous research that showed the central role of structural and institutional barriers in shaping health-seeking behaviors in low-resources settings, our findings showed that exposure to armed conflict and chronic insecurity not only increased these barriers but also reshaped caregiverś perceptions of trust and risk in ways that directly contributed to vaccine hesitancy. Unlike studies that focus solely on access to services or health literacy as explanatory factors, our study highlighted how socio-political instability intersected with community-level dynamics, such as gender roles and informal social networks, to shape behaviours in complex and sometimes contradictory ways. Both our study and the work of Ager et al. (2015) emphasised the critical importance of formal and informal partnerships in building systemic resilience. These partnerships are especially vital in conflict-affected regions where conventional health systems infrastructures are often compromised or entirely absent.

Our findings have several implications for policy and practice. First, rebuilding trust in health systems requires culturally sensitive interventions that address the specific needs and concerns of caregivers. Strategies should include engaging community members, improving health infrastructure and ensuring consistent and reliable service delivery. Second, addressing economic barriers to provide financial support and improve access to care, thus, reducing inequalities. Third, strengthening health communication efforts, particularly in underserved areas, is critical to counter misinformation and promote positive health behaviours.

However, this qualitative study has several limitations that should be acknowledged. First, although qualitative approach allowed for in-depth exploration of caregivers’ experiences and provided rich contextual detail, the data collected in Borno, Yobe and Adamawa states may not fully represent experiences in all conflict-affected regions of northeast Nigeria or reflect the heterogeneity of experiences across communities. Second, the use of focus group discussions may have introduced social desirability bias or pressure to conform, potentially limiting the openness or diversity of views expressed by participants. Third, data collection took place in communities experiencing ongoing armed violence, which may have influenced participants’ responses – the sensitive security context and perceived risks may have prevented participants from fully disclosing experiences of violence, mistrust or negative perceptions of health services, potentially resulting in incomplete narratives.

## Conclusion

This study provides critical insights into how intersecting factors - such as health-seeking behaviour, trust in health systems, exposure to conflict and vaccine hesitancy - shape caregivers’ decisions to seek health services for their children in conflict-affected northeast Nigeria. The findings highlight that insecurity, economic hardship, misinformation, patriarchal decision-making structures and widespread mistrust of health systems combine to create significant barriers to timely health care seeking, including routine immunisation. Despite these significant challenges, caregivers demonstrate resilience and resourcefulness, developing informal, community-based strategies to overcome barriers to health care and mitigate risks.

## Data Availability

All data produced in the present study are available upon reasonable request to the authors

